# Risk Factors of Multi-Drug Resistant Tuberculosis among Tuberculosis Patients in Province 3, Nepal: A case-control study

**DOI:** 10.1101/2024.06.16.24309002

**Authors:** Puspa Acharya, Niraj Bhattarai, Bhuban Raj Kunwar, Khem Raj Sharma, Vijay Kumar Khanal, Birendra Kumar Yadav

## Abstract

**Background:** Drug-resistant tuberculosis poses a significant threat to global TB control efforts, potentially reversing progress made in reducing TB-related morbidity and mortality. This study aims to identify risk factors for multidrug-resistant TB [MDR-TB] in Province 3, Nepal.

**Methodology:** A case-control study was conducted by matching TB-infected patients undergoing MDR-TB treatment and DS-TB treatment by gender. Data was collected through structured questionnaires and interviews and analyzed using binary logistic regression.

**Results:** Significant risk factors for MDR-TB included Pulmonary Tuberculosis [PTB], previous TB treatment history, close contact with DR-TB patients, and subjective feelings of sadness.

**Conclusion:** The study highlights the importance of prevention measures to break transmission chains and infection control in health facilities. Additionally, it underscores the need for mental health support for TB patients.

## Introduction

Tuberculosis [TB] is the most common cause of death worldwide and the leading cause of death from a single infectious agent, even exceeding HIV/AIDS. An estimated 10.0 million [range, 9.0– 11.1 million] people fell ill with TB in 2018; there were 1.2 million TB deaths among HIV-negative people in 2018 and a further 2,51,000 deaths among HIV-positive people(1).In April 1993, the World Health Organization [WHO] declared tuberculosis a public health emergency[2]. After defining the nature and size of the global TB problem, WHO introduced the Directly Observed Treatment Short-course [DOTS] as a solution to an emergency[3]. The expanding HIV epidemic and the growth of drug-resistant tuberculosis [DR-TB] further subverted the DOTS strategy, which was hampered by imprecise diagnostic tools and passive case detection(4).

Antimicrobial Resistance [AMR] is an increasingly significant threat to public health, and the rise of AMR coincides with the development of TB (5,6). The rise of drug-resistant tuberculosis has the potential to reverse progress made to scale back tuberculosis-related morbidity and mortality over the past 20 years. Drug resistance emerges as a result of inadequate tuberculosis treatment, which could be an incorrect combination of tuberculosis drugs, inadequate dose or duration, or irregular drug-taking. These problems can occur in any setting but are particularly prevalent in poorly regulated non-public sectors(7). Multi-drug resistant tuberculosis [MDR-TB] is tuberculosis that is resistant to at least two first-line drugs, isoniazid and rifampicin(8).TB strains with DR-TB are harder to treat than drug-susceptible ones and also take much longer(9). Globally, only half of the MDR-TB patients are treated successfully; the other half are at high risk of either not surviving or continuing to transmit the disease and threaten global progress towards the targets set by the End TB Strategy of the World Health Organization [WHO](4). Globally, in 2018, an estimated 3.4% [95% confidence interval [CI]: 2.5–4.4%] of new cases and 18% [95% CI: 7.6–31%] of previously treated cases had MDR/RR-TB. There were 484,000 [range, 417,000–556,000] incident cases of MDR/RR-TB in 2018, and only 32% of the estimated patients were enrolled in treatment. The highest proportions are in several countries of the former Soviet Union [above 25% in new cases and above 50% in previously treated cases. World Health Organization [WHO] has recently updated the guidance on DR-TB treatment by including Levofloxacin [a fluoroquinolone] and pyrazinamide as a component of the treatment regimens for MDR/RR-TB and isoniazid-resistant, rifampicin-susceptible TB [Hr-TB](1). As MDR-TB involves more extensive and longer treatment courses than drug-susceptible TB, households with multidrug resistances are at a particularly greater risk of incurring catastrophic costs(10). The percentage facing catastrophic total costs ranged from 27% to 83% for all forms of TB and from 67% to 100% for drug-resistant TB (1). The disease can affect anyone anywhere, but most people who develop TB [about 90%] are adults, and the male: female ratio is 2:1[1]; long-term exposure to ambient SO2 is likely to increase the risk of TB in males(11). Global targets and milestones for reducing the burden of TB disease have been set as part of the Sustainable Development Goals [SDGs] and the World Health Organization’s End TB Strategy, and seven countries with high TB burdens were also on track to achieve 2020 END TB milestones during this period (1).

Tuberculosis [TB] remains a serious public health problem in the South-East Asia Region [SEAR] as well (12). DR-TB burden was higher in India [27%], China [14%] and the Russian Federation [9%] in 2018(1).TB is also the leading cause of disability-adjusted life years [DALYs] lost among people in the region among all communicable diseases, affecting the productive age group of the countries in SEAR(13). Similarly, Myanmar has over 70% of TSR for MDR-TB and was on track to achieve 2020 END TB milestones.(1).

It has been predicted that deaths attributable to AMR could rise more than tenfold to 10 million annually by 2050. Unless action is taken now, DR-TB could be responsible for about 2.5 million of these deaths(14). A global total of 186 772 cases of multidrug-resistant TB or rifampicin-resistant TB [MDR/RR-TB] were notified in 2018, up from 160 684 in 2017, and 156 071 cases were enrolled in treatment, up from 139 114 in 2017. Despite these improvements, only 32% of the estimated 484,000 cases [range 417,000–556,000] were enrolled in treatment in 2018(1). The uncontrolled TB burden impacts socio-economic development and increases drug resistance in the SEAR region (15). If tuberculosis management practices across sectors in India remain unchanged over the next 20 years, it is estimated a 47% increase in the incidence of isoniazid resistance, a 152% increase in MDR-TB incidence, a 242% increase in prevalent untreated multidrug-resistant tuberculosis, and a 275% increase in the risk of multidrug-resistant tuberculosis infection(7).

In Nepal, there are approximately 1500 [0.84 to 2.4] cases of DR-TB annually, and only 350 to 450 MDR-TB cases are reported annually. So, the missing cases are the major problem in Nepal(8). There are major challenges for diagnosis of MDR TB in Nepal, such as inadequate training, frequent power failure, difficulty in maintaining an appropriate steady temperature, module failure, which is often not replaced in time, issues with calibration and timely availability of cartridges, as well as appropriate ways to store the new cartridges and safe disposal of the used cartridges (16). From a public health perspective, the MDR-TB growing epidemics will not be controlled merely by the introduction of a few new antibiotics as it is also linked to patient compliance and adequate case management supported by efficient TB programs(17). Deaths from drug-resistant TB now account for about one-third of all antimicrobial resistance deaths worldwide. Treating DR-TB is costlier and may take three to fourfold as long as not all people will survive.(6). The latest treatment outcome data for people with MDR/RR-TB showed a global treatment success rate of only 56%(1).

As MDR-TB involves more extensive and longer treatment courses than drug-susceptible TB, households with multidrug resistance are at a particularly greater risk of incurring catastrophic costs. The study identified the factors associated with resistance that might assist concerned authorities in focusing efforts on those factors in the community.This might help prevent and control tuberculosis and MDR-TB by influencing Public Health Policy. As MDR-TB involves more extensive and longer treatment courses than drug-susceptible TB, households with multidrug resistance are at a particularly greater risk of incurring catastrophic costs. Hence, ending TB will benefit the poorest ones.(10). This study aims to assess the risk factors of Multi-Drug-Resistant Tuberculosis among Tuberculosis patients of Province 3, Nepal.

## METHODS

The study used a case-control design to evaluate the risk of MDR-TB, which was conducted in Province 3, Nepal, which is known for its high MDR-TB burden. Cases comprised confirmed MDR-TB patients undergoing treatment at selected MDR treatment centres and sub-centres in Kathmandu, Bhaktapur, and Chitwan. At the same time, controls consisted of TB patients receiving anti-tuberculosis drugs at designated DOTS centres in the same districts. Inclusion criteria for cases required confirmation of MDR-TB through records from the MDR-TB treatment center, while controls encompassed pulmonary and extra-pulmonary TB patients on anti-tuberculosis medication. Exclusions were applied to severely ill patients who were unable to participate and those under 15 years old. The study adhered to ethical considerations, received clearance from the institutional review committee, and obtained informed verbal consent from respondents, ensuring confidentiality and data integrity. The researcher also took permission from the National Tuberculosis Control Centre, Thimi Bhaktapur, to conduct this research in the selected study sites after receiving IRC clearance. The structured questionnaire was prepared after a substantial literature review, and a piloting study was done among TB patients of BPKIHs, Dharan, by obtaining verbal consent from the participants. The study was conducted from March 2021 to August 2021. The operational definition used in the study is included as a supplementary document.

### Sampling Technique

The three districts were selected randomly, and all the DR-TB centres from these districts were included in the study. The subjects who gave consent were interviewed. Cases were selected from these DR-TB treatment centres/subcentre such as Nepal Anti-TB Association-Kathmandu, Japan Nepal Tuberculosis Research association-Kathmandu, National Tuberculosis Control centre-Bhaktapur, Nepal Anti-TB Association-Chitwan. All the DS-TB cases with approved verbal consent were interviewed. DOTS centres such as Manmohan Memorial Medical College and Teaching Hospital-Kathmandu, Nagarik Community Hospital-Bhaktapur, Bharatpur Health office, Parbatipur Health Post, Genetup-Kathmandu, JANTRA-Kathmandu, NATA-Chitwan were selected for this study.

### Calculation of the sample size

The study period was fixed for six months, and during the study periods, all the laboratory-confirmed and registered DR-TB patients were asked to participate from all treatment centres/sub-centres in selected districts—Chitwan, Kathmandu, and Bhaktapur. Despite approximately 85 DR-TB patients across these districts, only 77 agreed to participate. All the DS-TB cases were also interviewed, and gender matching was ensured using SPSS, resulting in a final sample size of 154 respondents, maintaining a 1:1 ratio of cases to controls.

### Data Collection Techniques

Amidst the challenges posed by the COVID-19 pandemic, a second wave in Nepal, a face-to-face interview was undertaken to collect data from both cases and controls. The clinical attributes of the participants were meticulously validated against their laboratory records. Despite the pandemic’s constraints, conducting interviews proved to be difficult. However, the institution provided a designated counselling room, coupled with the use of appropriate personal protective equipment [PPE] and adherence to social distancing measures; data collection was successfully facilitated. Recent episodes of TB history were taken to avoid recall bias, and a researcher collected data herself to minimise interviewer variations.

### Data Management and Analysis

Data entry was conducted using Microsoft Excel and subsequently transferred to SPSS version 16 for statistical analysis. Before entry, numerical coding was manually prepared based on the nature of the data, and regular cross-checking ensured accuracy, with verification after every ten entries. Descriptive statistics, Frequency, and percentages were employed to present case and control frequencies. The inferential analysis utilised chi-square tests for categorical nominal data association and binary regression analysis for variables potentially predictive of MDR-TB, with significance set at p < 0.05 and 95% confidence intervals. Factors exhibiting a p-value less than 0.020 in bivariate analysis underwent binary logistic regression for further analysis after adjusting confounders.

### Variables of the study

#### Independent variable

✓ Age
✓ Gender
✓ Weight
✓ Marital status
✓ Residence
✓ Education
✓ Occupation
✓ Monthly income
✓ Ethnicity
✓ Religion
✓ Family type
✓ Co-Morbid (HIV/AIDS & Other diseases)
✓ stigma
✓ social isolation
✓ Means of transportation
✓ Subjective feelings of sadness

#### Associated factors

✓ Types of TB
✓ Previous History of TB
✓ Interruption of TB treatment
✓ Contact with TB patients
✓ Contact with DR TB patients
✓ Response to Treatment Phase
✓ Counselling session

#### Dependent variable

✓ Multi Drug Resistant Tuberculosis (MDR-TB)

## RESULTS

Table 1 illustrates demographic and socio-economic characteristics between cases and controls. While both groups have similar gender distributions, with males representing 55.8% and females 44.2%, the control group has a higher proportion of individuals under 35 years [64.9%] compared to the case group [58.4%]. Marital status and ethnicity are largely comparable, but there are differences in residence, education, and employment status. The control group has more residents in metropolitan cities [62.3% vs. 41.6% in cases] and fewer in rural areas [13.0% vs. 37.7% in cases]. Additionally, a higher percentage of cases are unemployed [44.2% vs. 33.8% in controls] and more are students [24.7% vs. 31.2% in controls]. Access to health facilities within 30 minutes is higher in the control group [96.1% vs. 66.2% in cases], and they utilize private health facilities more [85.7% vs. 58.8% in cases].

**Table 1:** Demographic and Socio-economic characteristics of respondents.

| Characteristics | Case [n=77]<br>Frequency [n] | Percentage<br>[%] | Control [n=77]<br>Frequency [n] | Percentage<br>[%] |
| --- | --- | --- | --- | --- |
| **Age** |  |  |  |  |
| < 35 | 45 | [58.4%] | 50 | [64.9%] |
| ≥ 35 | 32 | [41.6%] | 27 | [35.1%] |
| **Gender** |  |  |  |  |
| Male | 43 | [55.8%] | 43 | [55.8%] |
| Female | 34 | [44.2%] | 34 | [44.2%] |
| **Marital status** |  |  |  |  |
| Unmarried | 32 | [41.60%] | 35 | [45.50%] |
| Married | 43 | [55.80%] | 41 | [53.20%] |
| Widow | 1 | [1.30%] | 1 | [1.30%] |
| Widower | 1 | [1.30%] | 0 | [0.00%] |
| **Residence** |  |  |  |  |
| Metropolitan city | 32 | [41.6%] | 48 | [62.3%] |
| Municipality | 16 | [20.8%] | 19 | [24.7%] |
| Rural Municipality | 29 | [37.7%] | 10 | [13.0%] |
| **Education** |  |  |  |  |
| Uneducated | 5 | [6.5%] | 5 | [6.5%] |
| Primary level | 22 | [28.6%] | 13 | [16.9%] |
| Secondary level | 20 | [26.0%] | 28 | [36.4%] |
| Higher level education | 30 | [39.0%] | 31 | [40.3%] |
| **Occupation** |  |  |  |  |
| Unemployed | 34 | [44.2%] | 26 | [33.8%] |
| Formal sector employee | 10 | [13.0%] | 15 | [19.5%] |
| Informal sector employee | 14 | [18.2%] | 12 | [15.6%] |
| Student | 19 | [24.7%] | 24 | [31.2%] |
| **Monthly Income of employed respondents [n= 51]** |  |  |  |  |
| < Rs 24000 | 12 | [50%] | 15 | [55.6%] |
| ≥ Rs 24000 | 12 | [50%] | 12 | [44.4%] |
| **Ethnicity** |  |  |  |  |
| Dalit | 2 | [2.6%] | 4 | [5.2%] |
| Janajati | 41 | [53.2%] | 41 | [53.2%] |
| Madhesi | 2 | [2.6%] | 1 | [1.3%] |
| Muslim | 3 | [3.9%] | 1 | [1.3%] |
| Brahmin/Chhetri | 29 | [37.7%] | 29 | [37.7%] |
| Others | 0 | [0.0%] | 1 | [1.3%] |
| **Religion** |  |  |  |  |
| Hindu | 57 | [74.0%] | 65 | [84.4%] |
| Buddhist | 17 | [22.1%] | 8 | [10.4%] |
| Muslim | 3 | [3.9%] | 1 | [1.3%] |
| Christian | 0 | [0.0%] | 3 | [3.9%] |
| **Family Type** |  |  |  |  |
| Nuclear | 39 | [50.60%] | 46 | [59.70%] |
| Joint | 36 | [46.80%] | 30 | [39.00%] |
| Extended | 2 | [2.60%] | 1 | [1.30%] |
| **Presence of Adequate Ventilation** |  |  |  |  |
| No | 20 | [25.9%] | 8 | [10.3%] |
| Yes | 57 | [74.1%] | 69 | [89.7%] |
| **Recent Migration** |  |  |  |  |
| No | 35 | [45.5%] | 50 | [64.9%] |
| Yes | 42 | [54.5%] | 27 | [35.1%] |
| **Types of Migration** |  |  |  |  |
| Internal Migration | 37 | [88.1%] | 23 | [85.2%] |
| External Migration | 5 | [11.9%] | 4 | [14.8%] |
| **Time to reach Health Facilities** |  |  |  |  |
| Less than 30 minutes | 51 | [66.2%] | 74 | [96.1%] |
| Greater than 30 minutes | 26 | [33.8%] | 3 | [3.9%] |
| **Means of Transportation** |  |  |  |  |
| Cycle | 1 | [1.3%] | 2 | [2.6%] |
| Bike/Scooter | 11 | [14.3%] | 25 | [32.5%] |
| Public Vehicle | 64 | [83.1%] | 41 | [53.2%] |
| By foot | 1 | [1.3%] | 9 | [11.7%] |
| **Health-seeking Behavior of patients** |  |  |  |  |
| Too late | 57 | [74.0%] | 51 | [66.2%] |
| In time | 17 | [22.1%] | 25 | [32.5%] |
| Early | 3 | [3.9%] | 1 | [1.3%] |
| **Treatment Initiation site** |  |  |  |  |
| Public Health Facilities | 43 | [58.8%] | 11 | [14.3%] |
| Private Health Facilities | 34 | [42.2%] | 66 | [85.7%] |

Table 2 shows that pulmonary TB cases were higher among the cases, at 89.6%, compared to 57.1% in the control group. Conversely, Extra Pulmonary TB cases were more prevalent in the Controls at 42.9% versus 10.4% in the Cases. Notably, a prior history of TB was significantly more common among Cases, with 63.6% compared to a mere 10.4% in Controls. Among those with a TB history, Cases experienced treatment interruptions at a higher rate of 51.0%, contrasting with just 12.5% in Controls. Reasons for such interruptions varied, with complications accounting for 8% among Cases and none among Controls, while loss to follow-up stood at 16% for Cases versus 100% for Controls. Additionally, 25% of Cases switched to MDR treatment compared to none in Controls. Close contact with TB patients was markedly more prevalent among Cases [64.9%] than Controls [32.5%], with similar trends observed for contact with DR TB patients. Notably, negative sputum conversion rates were higher among Cases [79.2%] versus Controls [66.2%]. Hospital stays were significantly more common among Cases [89.6%] than Controls [31.2%]. Treatment modalities also exhibited disparities, with 44.2% of Cases receiving treatment after failure compared to none in Controls and 48.1% of Cases undergoing the intensive treatment phase versus 74.0% of Controls. In terms of TB treatment categories, Cases predominantly fell into LR1 [58.5%] and LR2 [31.1%] categories, while Controls were exclusively categorized as CAT I [100%].

**Table 2:** Clinical Characteristics of Respondents.

| Characteristics | Case[n=77]<br>Frequency | Percentage [%] | Control[n=77]<br>Frequency | Percentage [%] |
| --- | --- | --- | --- | --- |
| **Tuberculosis Types** |  |  |  |  |
| Pulmonary TB | 69 | [89.6%] | 44 | [57.1%] |
| Extra Pulmonary TB | 8 | [10.4%] | 33 | [42.9%] |
| **Previous history of TB** |  |  |  |  |
| No | 28 | [36.4%] | 69 | [89.6%] |
| Yes | 49 | [63.6%] | 8 | [10.4%] |
| **If yes, was there any interruption of TB treatment [n= 49 cases, 8 controls]** |  |  |  |  |
| No | 24 | [49.0%] | 7 | [87.5%] |
| Yes | 25 | [51.0%] | 1 | [12.5%] |
| **Reason for interruption of TB treatment [n= 25 cases, 1 controls]** |  |  |  |  |
| Complication | 2 | [8%] | 0 | [0%] |
| Loss to follow up | 4 | [16%] | 1 | [100%] |
| Switch to MDR | 5 | [25%] | 0 | [0%] |
| Site Changed | 1 | [4%] | 0 | [0%] |
| TAF | 12 | [48%] | 0 | [0%] |
| Workload | 1 | [4%] | 0 | [0%] |
| **Close contact with TB Patients** |  |  |  |  |
| No | 27 | [35.1%] | 52 | [67.5%] |
| Yes | 50 | [64.9%] | 25 | [32.5%] |
| **Close contact with DR TB patients** |  |  |  |  |
| No | 38 | [49.4%] | 76 | [98.7%] |
| Yes | 39 | [50.6%] | 1 | [1.3%] |
| **Response to TB treatment [Sputum Conversion]** |  |  |  |  |
| Negative | 61 | [79.2%] | 51 | [66.2%] |
| Positive | 16 | [20.8%] | 26 | [33.8%] |
| **Experience of hospital stays** |  |  |  |  |
| No | 8 | [10.4%] | 53 | [68.8%] |
| Yes | 69 | [89.6%] | 24 | [31.2%] |
| **Registration category of patients** |  |  |  |  |
| New | 23 | [29.9%] | 69 | [89.6%] |
| Relapse | 26 | [20.8%] | 8 | [10.4%] |
| Treatment After |  |  |  |  |
| Failure | 34 | [44.2%] | 0 | [0.0%] |
| Treatment after loss to follow-up | 2 | [2.6%] | 0 | [0.0%] |
| Others previously Treated | 1 | [1.3%] | 0 | [0.0%] |
| Previous Treatment History Unknown | 1 | [1.3%] | 0 | [0.0%] |
| **Treatment Phase** |  |  |  |  |
| Intensive | 37 | [48.1%] | 57 | [74.0%] |
| Continuous | 40 | [51.9%] | 20 | [26.0%] |
| **TB Treatment Category** |  |  |  |  |
| LR1 | 45 | [58.5%] | 0 | [0%] |
| LR2 | 24 | [31.1%] | 0 | [0%] |
| LR3 | 1 | [1.3%] | 0 | [0%] |
| SSTR | 4 | [5.2%] | 0 | [0%] |
| Others | 3 | [3.9%] | 0 | [0%] |
| CAT I | 0 | [0%] | 77 | [100%] |

Table 3 presents psychological and social tuberculosis-related variables [TB] among cases and controls. In the cases group, 54.5% attended TB counselling sessions, which was notably higher than 85.7% in the control group. Regarding HIV status, 93.5% of Cases were negative, compared to 100% of Controls. Additionally, 31.2% of Cases reported having other diseases, contrasting with 24.7% in the control group. Alcohol consumption was notably prevalent among Cases at 76.6%, whereas 68.8% of Controls reported the same. Smoking was more common among Cases [51.9%] than in Controls [18.2%], and similarly, illicit drug use was higher in Cases [18.2%] than in Controls [2.6%]. The incidence of imprisonment was minimal in both groups, with Cases slightly higher at 3.9% compared to Controls at 2.6%. Social isolation was more pronounced among Cases at 53.2% compared to Controls at 39.0%. Cases also reported higher rates of perceived stigma associated with TB, with 74.0% fearing loss of friends and employment, 77.9% experiencing social stigma, and 55.8% preferring privacy regarding their TB status, whereas Controls reported lower rates in these categories. Furthermore, Cases exhibited higher nervousness levels [76.6%] and sadness [58.4%] than Controls. Troubles within the past 12 months were reported by 46.8% of Cases and 14.3% of Controls. Despite challenges, both groups expressed happiness, with 79.2% of Cases and 88.3% of Controls considering themselves happy despite their TB status.

**Table 3:** Psychological and co-morbidity status of respondents.

| Variables | Case [n=77]<br>Frequency [n] | Percentage<br>[%] | Control [n=77]<br>Frequency [n] | Percentage<br>[%] |
| --- | --- | --- | --- | --- |
| **Attended any TB counselling sessions** |  |  |  |  |
| No | 35 | [45.50%] | 11 | [14.30%] |
| Yes | 42 | [54.50%] | 66 | [85.70%] |
| **HIV status** |  |  |  |  |
| Negative | 72 | [93.5%] | 77 | [100.0%] |
| Positive | 5 | [6.5%] | 0 | [00.0%] |
| **Presence of other disease** |  |  |  |  |
| No | 53 | [68.8%] | 58 | [75.3%] |
| Yes | 24 | [31.2%] | 19 | [24.7%] |
| **Alcohol consumption** |  |  |  |  |
| No | 18 | [23.4%] | 24 | [31.2%] |
| Yes | 59 | [76.6%] | 53 | [68.8%] |
| **Smoking** |  |  |  |  |
| No | 37 | [48.1%] | 63 | [81.8%] |
| Yes | 40 | [51.9%] | 14 | [18.2%] |
| **Illicit drug use** |  |  |  |  |
| No | 63 | [81.8%] | 75 | [97.4%] |
| Yes | 14 | [18.2%] | 2 | [2.6%] |
| **Imprisonment** |  |  |  |  |
| No | 74 | [96.10%] | 75 | [97.40%] |
| yes | 3 | 3 [3.90%] | 2 | [2.60%] |
| **Social isolation** |  |  |  |  |
| No | 36 | [46.8%] | 30 | [39.0%] |
| Yes | 41 | [53.2%] | 47 | [61.0%] |
| **I feel that I will lose friends and employment because I have TB** |  |  |  |  |
| No | 20 | [26.0%] | 55 | [71.4%] |
| Yes | 57 | [74.0%] | 22 | [28.6%] |
| **People look at me differently because I have tuberculosis** |  |  |  |  |
| No | 17 | [22.1%] | 49 | [63.6%] |
| Yes | 60 | [77.9%] | 28 | [36.4%] |
| **I don't like other people to know that I have TB** |  |  |  |  |
| No | 34 | [44.2%] | 24 | [31.2%] |
| Yes | 43 | [55.8%] | 53 | [68.8%] |
| **I consider myself nervous** |  |  |  |  |
| No | 18 | [23.4%] | 41 | [53.2%] |
| Yes | 59 | [76.6%] | 36 | [46.8%] |
| **Sometimes I feel so sad that nothing can cheer me up** |  |  |  |  |
| No | 32 | [41.6%] | 68 | [88.3%] |
| Yes | 45 | [58.4%] | 9 | [11.7%] |
| **Did something trouble you during the past 12 months?** |  |  |  |  |
| No | 41 | [53.2%] | 66 | [85.7%] |
| Yes | 36 | [46.8%] | 11 | [14.3%] |
| **Despite of disease, I think I am happy.** |  |  |  |  |
| No | 16 | [20.80%] | 16 | [11.70%] |
| Yes | 61 | [79.20%] | 61 | [88.30%] |

Table 4 shows several significant associations highlighting key environmental and socio-economic factors. Cases group exhibit a notably higher prevalence in rural areas, with 37.7% residing in rural municipalities compared to only 13.0% of controls [p = 0.02]. Recent migration emerges as a significant factor, with 54.5% of Cases reporting recent migration compared to 35.1% of controls [p = 0.015]. Additionally, inadequate ventilation is more prevalent among the cases group, with 26.0% reporting inadequate ventilation compared to 10.4% of controls [p = 0.012]. Access to healthcare services is also impacted, as evidenced by 66.2% of cases taking more than 30 minutes to reach health facilities compared to only 3.9% of controls [p < 0.001]. These findings underscore the critical influence of environmental and socio-economic determinants on TB prevalence and access to healthcare, emphasizing the importance of targeted interventions to address these disparities for effective TB prevention and control strategies.

**Table 4.**
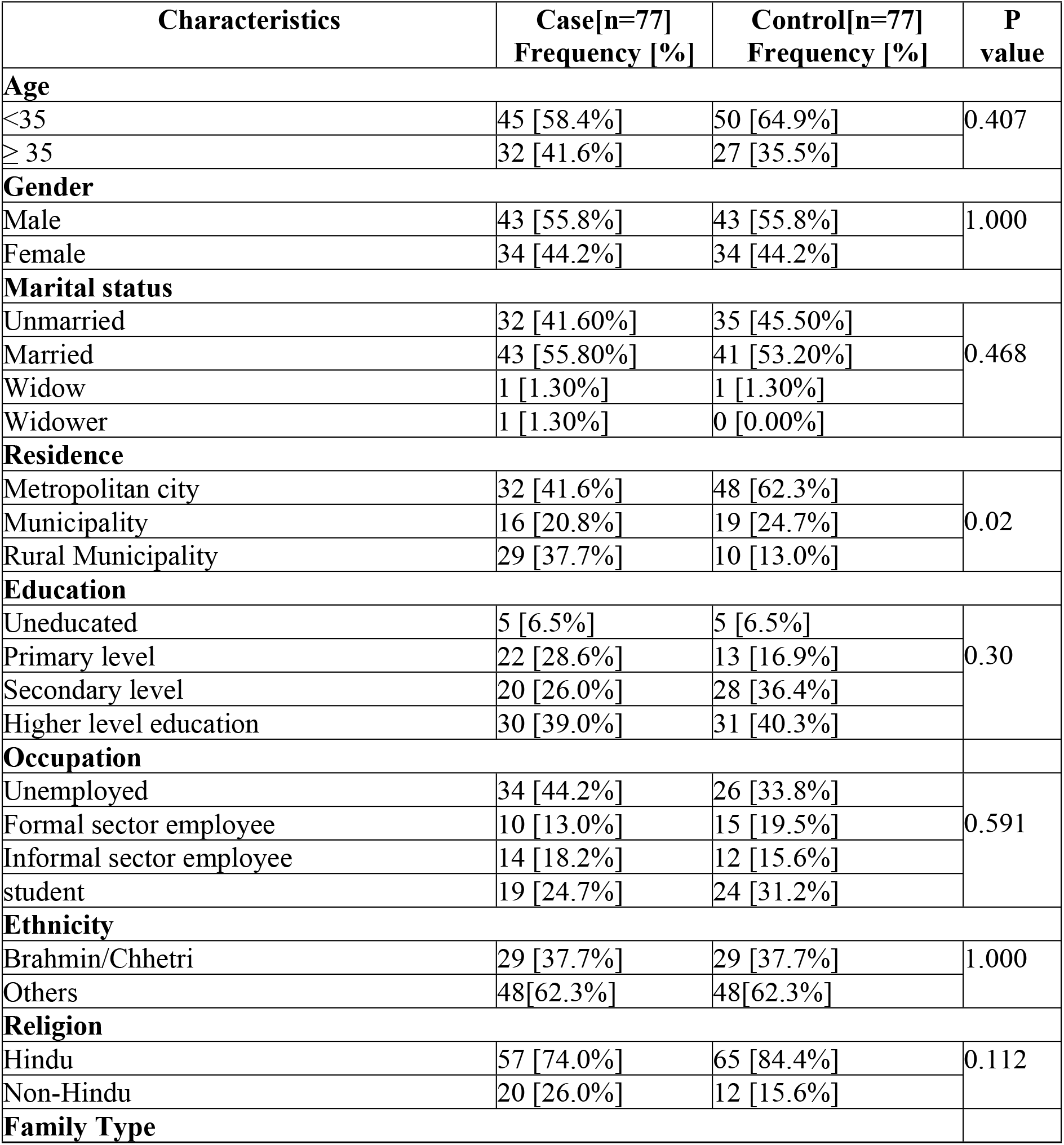
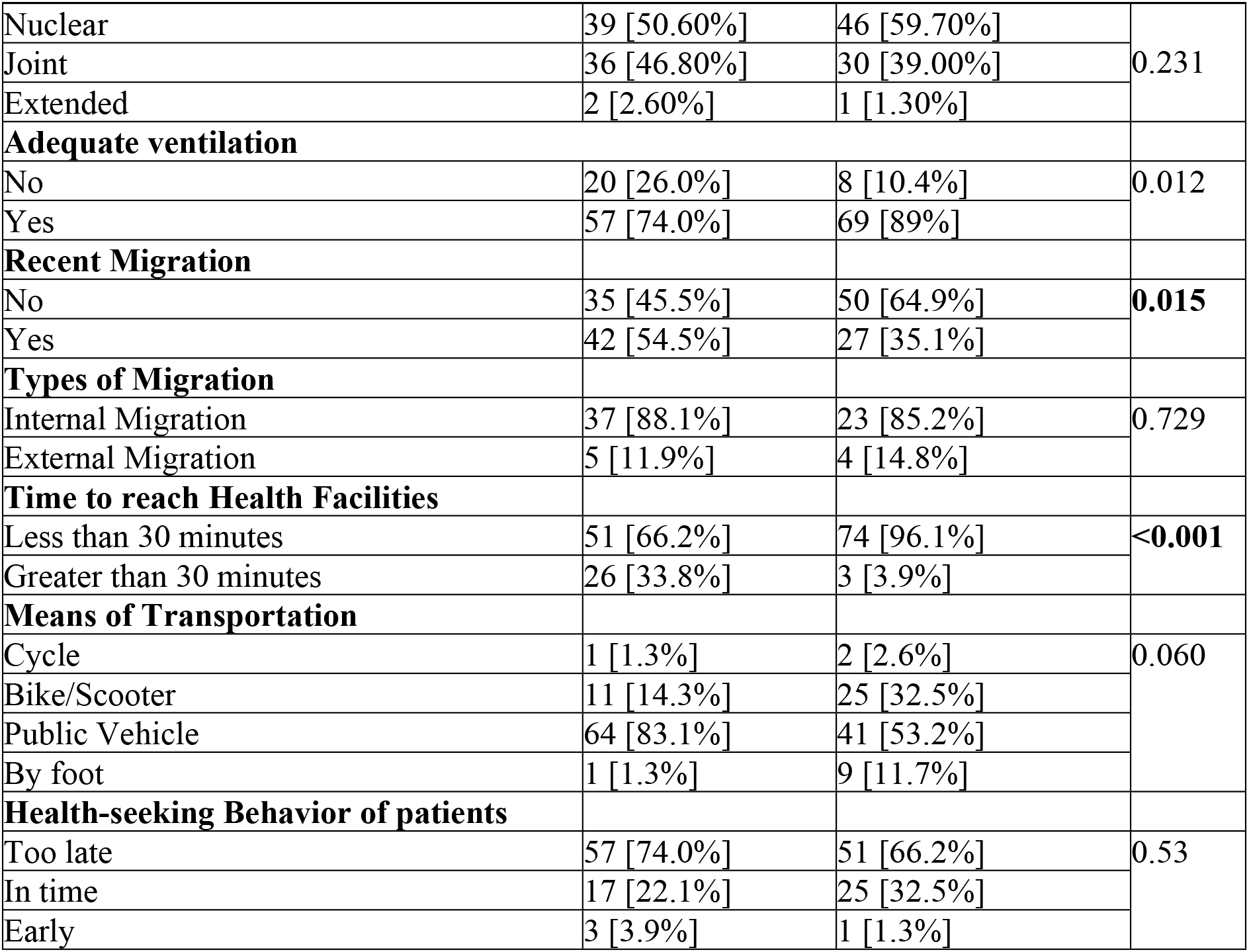
: Key Factors associated with MDR-TB.

| Characteristics | Case[n=77]<br>Frequency [%] | Control[n=77]<br>Frequency [%] | P<br>value |
| --- | --- | --- | --- |
| Age |  |  |  |
| <35 | 45 [58.4%] | 50 [64.9%] | 0.407 |
| ≥ 35 | 32 [41.6%] | 27 [35.5%] |  |
| Gender |  |  |  |
| Male | 43 [55.8%] | 43 [55.8%] | 1.000 |
| Female | 34 [44.2%] | 34 [44.2%] |  |
| Marital status |  |  |  |
| Unmarried | 32 [41.60%] | 35 [45.50%] | 0.468 |
| Married | 43 [55.80%] | 41 [53.20%] |  |
| Widow | 1 [1.30%] | 1 [1.30%] |  |
| Widower | 1 [1.30%] | 0 [0.00%] |  |
| Residence |  |  |  |
| Metropolitan city | 32 [41.6%] | 48 [62.3%] | 0.02 |
| Municipality | 16 [20.8%] | 19 [24.7%] |  |
| Rural Municipality | 29 [37.7%] | 10 [13.0%] |  |
| Education |  |  |  |
| Uneducated | 5 [6.5%] | 5 [6.5%] | 0.30 |
| Primary level | 22 [28.6%] | 13 [16.9%] |  |
| Secondary level | 20 [26.0%] | 28 [36.4%] |  |
| Higher level education | 30 [39.0%] | 31 [40.3%] |  |
| Occupation |  |  |  |
| Unemployed | 34 [44.2%] | 26 [33.8%] | 0.591 |
| Formal sector employee | 10 [13.0%] | 15 [19.5%] |  |
| Informal sector employee | 14 [18.2%] | 12 [15.6%] |  |
| student | 19 [24.7%] | 24 [31.2%] |  |
| Ethnicity |  |  |  |
| Brahmin/Chhetri | 29 [37.7%] | 29 [37.7%] | 1.000 |
| Others | 48[62.3%] | 48[62.3%] |  |
| Religion |  |  |  |
| Hindu | 57 [74.0%] | 65 [84.4%] | 0.112 |
| Non-Hindu | 20 [26.0%] | 12 [15.6%] |  |
| Family Type |  |  |  |
| Nuclear | 39 [50.60%] | 46 [59.70%] | 0.231 |
| Joint | 36 [46.80%] | 30 [39.00%] |  |
| Extended | 2 [2.60%] | 1 [1.30%] |  |
| **Adequate ventilation** |  |  |  |
| No | 20 [26.0%] | 8 [10.4%] | 0.012 |
| Yes | 57 [74.0%] | 69 [89%] |  |
| **Recent Migration** |  |  |  |
| No | 35 [45.5%] | 50 [64.9%] | **0.015** |
| Yes | 42 [54.5%] | 27 [35.1%] |  |
| **Types of Migration** |  |  |  |
| Internal Migration | 37 [88.1%] | 23 [85.2%] | 0.729 |
| External Migration | 5 [11.9%] | 4 [14.8%] |  |
| **Time to reach Health Facilities** |  |  |  |
| Less than 30 minutes | 51 [66.2%] | 74 [96.1%] | **<0.001** |
| Greater than 30 minutes | 26 [33.8%] | 3 [3.9%] |  |
| **Means of Transportation** |  |  |  |
| Cycle | 1 [1.3%] | 2 [2.6%] | 0.060 |
| Bike/Scooter | 11 [14.3%] | 25 [32.5%] |  |
| Public Vehicle | 64 [83.1%] | 41 [53.2%] |  |
| By foot | 1 [1.3%] | 9 [11.7%] |  |
| **Health-seeking Behavior of patients** |  |  |  |
| Too late | 57 [74.0%] | 51 [66.2%] | 0.53 |
| In time | 17 [22.1%] | 25 [32.5%] |  |
| Early | 3 [3.9%] | 1 [1.3%] |  |

Table 5 depicted Pulmonary TB as notably more prevalent among cases [89.6%] compared to controls [57.1%], indicating it as a significant risk factor [p < 0.001; OR: 6.5; 95% CI: 2.7-15.2]. Conversely, controls exhibited a higher incidence of Extra Pulmonary TB [42.9%] compared to cases [10.4%]. Individuals with a previous TB history were notably more common among cases [63.6%] than controls [10.4%], indicating it as a substantial risk factor [p < 0.001; OR: 15.09; 95% CI: 6.34-35.91]. Similarly, cases were more likely to report close contact with TB patients [64.9%] compared to controls [32.5%], showing a significant association [p < 0.001; OR: 3.85; 95% CI: 1.97-7.51] likewise, cases were significantly associated with close contact with drug-resistant TB patients [p = 0.001; OR: 78; 95% CI: 10.31-589.62] suggesting the exposure as a risk factor. Cases reported higher levels of perceived stigma, with concerns about losing friends and employment due to TB [p < 0.001; OR: 7.12; 95% CI: 3.50-14.49] and feeling that people looked at them differently [p < 0.001; OR: 6.17; 95% CI: 3.02-12.57], indicating poor psychosocial health. Additionally, cases exhibited a higher prevalence of nervousness [p < 0.001; OR: 3.73; 95% CI: 1.86-7.45] and consistent sadness [p < 0.001; OR: 10.62; 95% CI: 4.63-24.36], signifying these as factors of concern. These findings highlight the importance of addressing these psychosocial risk factors alongside medical interventions in TB management.

**Table 5:** Association of MDR–TB with clinical characteristics of the respondent.

| Categories | Case[n=77] | Control [n=77] | P-value | OR | 95% CI<br>[Lower-Upper] |
| --- | --- | --- | --- | --- | --- |
| Tuberculosis Types |  |  |  |  |  |
| Pulmonary TB | 69 [89.6%] | 44 [57.1%] | <0.001 | 6.5 | 2.7-15.2 |
| Extra Pulmonary TB | 08 [10.4%] | 33 [42.9%] |  |  |  |
| Previous history of TB |  |  |  |  |  |
| Yes | 49 [63.6%] | 08 [10.4%] | <0.001 | 15.09 | 6.34-35.91 |
| No | 28 [36.4%] | 69 [89.6%] |  |  |  |
| If yes, was there any interruption of TB treatment |  |  |  |  |  |
| Yes | 25 [51.0%] | 01 [12.5%] | 0.059 | 7.29 | 0.83-63.70 |
| No | 24 [49.0%] | 07 [87.5%] |  |  |  |
| Close contact with TB Patients |  |  |  |  |  |
| Yes | 50 [64.9%] | 25 [32.5%] | <0.001 | 3.85 | 1.97-7.51 |
| No | 27 [35.1%] | 52 [67.5%] |  |  |  |
| Close contact with Drug Resistant TB patients |  |  |  |  |  |
| Yes | 39 [50.6%] | 01 [1.3%] | 0.001 | 78 | 10.31-589.62 |
| No | 38 [49.4%] | 76 [98.7%] |  |  |  |
| Response to TB treatment |  |  |  |  |  |
| Positive | 16 [20.8%] | 26 [33.8%] | 0.07 | 0.51 | 0.24 -1.06 |
| Negative | 61 [79.2%] | 51 [66.2%] |  |  |  |
| Experience of hospital stays |  |  |  |  |  |
| Yes | 69 [89.6%] | 24 [31.2%] | <0.001 | 19.04 | 7.92-45.76 |
| No | 08 [10.4%] | 53 [68.8%] |  |  |  |
| Attended any TB counselling sessions |  |  |  |  |  |
| Yes | 42 [54.50%] | 66 [85.70%] | <0.001 | 5 | 2.29-10.90 |
| No | 35 [45.50%] | 11 [14.30%] |  |  |  |
| HIV status |  |  |  |  |  |
| Positive | 5 [6.5%] | 0 [0.0%] | 0.02 | - |  |
| Negative | 72 [93.5%] | 77[100.0%] |  |  |  |
| Presence of other disease |  |  |  |  |  |
| Yes | 24 [31.2%] | 19 [24.7%] | 0.36 | 1.38 | 0.68-2.80 |
| No | 53 [68.8%] | 58 [75.3%] |  |  |  |
| Alcohol Consumption |  |  |  |  |  |
| Yes | 59 [76.6%] | 53 [68.8%] | 0.278 | 1.48 | 0.72-3.03 |
| No | 18 [23.4%] | 24 [31.2%] |  |  |  |
| Smoking |  |  |  |  |  |
| Yes | 40 [51.9%] | 14 [18.2%] | <0.001 | 4.86 | 2.34-10.11 |
| No | 37 [48.1%] | 63 [81.8%] |  |  |  |
| Illicit Drug |  |  |  |  |  |
| Yes | 14 [18.2%] | 2 [2.6%] | 0.002 | 8.33 | 1.82-38.06 |
| No | 63 [81.8%] | 75 [97.4%] |  |  |  |
| Imprisonment |  |  |  |  |  |
| Yes | 3 [3.90%] | 2 [2.60%] | 1.00 | 1.52 | 0.24-9.36 |
| No | 74 [96.10%] | 75 [97.40%] |  |  |  |
| **Social Isolation** |  |  |  |  |  |
| Yes | 41 [53.2%] | 47 [61.0%] | 0.329 | 0.72 | 0.38-1.37 |
| No | 36 [46.8%] | 30 [39.0%] |  |  |  |
| **I feel that I will lose friends and employment because I have TB** |  |  |  |  |  |
| Yes | 57 [74.0%] | 22 [28.6%] | <0.001 | 7.12 | 3.50-14.49 |
| No | 20 [26.0%] | 55 [71.4%] |  |  |  |
| **People look at me differently because I have tuberculosis.** |  |  |  |  |  |
| Yes | 60 [77.9%] | 28 [36.4%] | <0.001 | 6.17 | 3.02-12.57 |
| No | 17 [22.1%] | 49 [63.6%] |  |  |  |
| **I don't like other people to know that I have TB** |  |  |  |  |  |
| Yes | 43 [55.8%] | 53 [68.8%] | <0.001 | 0.57 | 0.29-1.10 |
| No | 34 [44.2%] | 24 [31.2%] |  |  |  |
| **I consider myself nervous.** |  |  |  |  |  |
| Yes | 59 [76.6%] | 36 [46.8%] | <0.001 | 3.73 | 1.86-7.45 |
| No | 18 [23.4%] | 41 [53.2%] |  |  |  |
| **Sometimes, I feel so sad that nothing can cheer me up** |  |  |  |  |  |
| Yes | 45 [58.4%] | 9 [11.7%] | <0.001 | 10.62 | 4.63-24.36 |
| No | 32 [41.6%] | 68 [88.3%] |  |  |  |
| **Did something trouble you during the past 12 months?** |  |  |  |  |  |
| Yes | 36 [46.8%] | 11 [14.3%] | <0.001 | 5.26 | 2.41-11.48 |
| No | 41 [53.2%] | 66 [85.7%] |  |  |  |
| **Despite of disease, I think I am happy.** |  |  |  |  |  |
| Yes | 61 [79.20%] | 68 [88.30%] | 0.126 | 1.98 | 0.81-4.81 |
| No | 16 [20.80%] | 9 [11.70%] |  |  |  |

After adjusting for the confounding factors, the final regression model [Table 6] showed that subjects infected with pulmonary tuberculosis were 14 times more at risk for MDR-TB than respondents with Extrapulmonary. Respondents who had previous TB treatment history were found to have more than 5-fold risk for MDR TB than those who were not previously treated. Similarly, respondents who were in close contact with DR-TB patients were found to be almost 22 times the risk for MDR-TB. In addition, MDR-TB patients were more [OR= 42.33, CI 1.36-1315.4] likely to perceive subjective feeling of sadness.

**Table 6.** Multivariate analysis of factors associated with MDR TB.

| Variables | Cat | $\beta$ | P value | Odds Ratio | 95% C.I. for OR | |
| --- | --- | --- | --- | --- | --- | --- |
|  |  |  |  |  | Lower | Upper |
| **Tuberculosis Type** | EPTB** | Ref. |  |  |  |  |
|  | PTB | 2.69 | 0.004 | 14.7 | 2.321 | 93.54 |
| **Previous TB treatment History** | No ** |  |  |  |  |  |
|  | Yes | 1.622 | 0.046 | 5.06 | 1.03 | 24.87 |
| **Close contact with DR TB patients** | No** |  |  |  |  |  |
|  | Yes | 3.099 | 0.015 | 22.18 | 1.829 | 269.1 |
| **Sometimes, I feel so sad that nothing can cheer me up** | No** |  |  |  |  |  |
|  | Yes | 3.746 | 0.033 | 42.35 | 1.364 | 1315.4 |
| **Constant** |  | -5.342 | 0.013 | 0.005 |  |  |

\*\* : Reference, Cat: Categories, N= 154

Cases [Y] = β0 + β1 [PTB]+ β2 [PH]+ β3 [DR-TB-C] + β4[sad] Y= [-5.342] + 2.6 + 1.622 + 3.099 + 3.746

Y= 5.725

This equation showed that one unit change in independent variables causes a 5.725 times change in dependent variables.

## Discussion

The study has provided relevant information about factors associated with MDR-TB, which can support activities being implemented to decrease the burden of TB in Nepal for policymakers. This study showed that Pulmonary Tuberculosis type, previous TB treatment history, close contact with DR-TB patients and subjective feelings of sadness were the strong predictors for MDR-TB.

Among sociodemographic factors, we identified that people residing in rural municipalities are more likely to develop MDR-TB than those living in urban areas [p=0.02], as supported by other studies(18,19). These data are alarming given the geographical features of our country like Nepal. Whereas the study conducted in Ukraine showed that the prevalence of RR/MDR-TB in the rural population was significantly lower than in the urban population at 20.9% and 29.0%, respectively [P = 0.000](20). This study also showed migration as one of the risk factors of MDR-TB [P = 0.015, OR = 2.22, 95% CI 1.16-4.25]. The study conducted in China showed that MDR is more prevalent among migrants [odds ratios [OR] 1.32, 95% confidence interval [CI] 1.02–1.72](21); migration is considered an important contributor to missing cases and may lead to poor outcomes(22). To mitigate the number of migrants, it’s essential to ensure uniform standards of care accessible nationwide, facilitated by robust communication channels such as telemedicine and referral systems. Additionally, implementing a unique identifier system would enable seamless patient connectivity across provinces and the entire nation.

This study showed that inadequate ventilation at home was a risk factor for MDR-TB [p = 0.01]. Poor ventilation and overcrowding have been documented as a risk factor for MDR-TB in several other studies in a variety of settings as well(23,24). Likewise, those subjects who lived in a house with no window or one window were almost two times more likely to develop airborne infection compared to people whose house has multiple windows [AOR = 1.81; 95% CI:1.06, 3.07](25), which may be due to the availability of favourable environment for MDR-TB transmission. Therefore, advocating for adequate ventilation is crucial to disrupt the transmission chain of MDR-TB.

Our study showed clinical features such as types of TB, previous TB treatment history, close contact with TB and DR-TB patients, and experience of hospital stays as the risk factors of MDR-TB with p = 0.001 for each factor. The majority of cases [89.6%]and controls [57.1%] had Pulmonary Tuberculosis, which is similar to the study conducted in Ethiopia, with the majority of participants having pulmonary TB in cases 141 [92.2%] and controls 137 [89.5%] (26). We found that patients having Pulmonary Tuberculosis were more likely to develop MDR-TB than those of Extra pulmonary TB patients, which is statistically significant in the final binary logistic regression model with OR = 14.735, 95% CI [2.321-93.54], as confirmed by the different studies in different countries; Germany(27), India(28) and Thailand(29).

This study showed previous TB treatment history was strongly associated with MDR-TB in final regression model, with OR= 5.601 [95% CI, 1.03-24.876]. The increased proportion of MDR-TB among previously treated cases has been indicated in various studies of various settings(28–33); The rise in MDR-TB cases among previously treated individuals underscores the urgency for higher quality treatment to prevent the development of drug resistance. Our study showed that, 51.0% of MDR cases had interruption of TB treatment, which is similar to the study conducted by Sanju B et al in 2017(34). We found patients with Interruption in TB treatment were more likely to develop MDR-TB than those without TB-Treatment interruption [p= 0.059, OR= 7.292, 95%CI 0.83-63.79], and another study also confirmed interruption of treatment as a risk factors of MDR-TB [28,35–38]. Interruption of TB treatment promotes the risk of bacterial mutations that eventually culminate in relapses and MDR-TB.

Our study showed that 64.9% of cases had history of contact with TB patient. Those who had history of close contact with TB patients were more likely to develop MDR-TB than those without close contact [p<0.001, OR=3.85, CI 1.974-71.5]. Several study conducted in different settings also confirmed that close contact with TB patients is a risk factors of MDR-TB[31,34,39], which is contrast to the study conducted in Ethiopia(40).

Similarly, Close contact with DR-TB patients was found to be a risk factor of MDR-TB. Final regression model indicated that close contact with DR-TB patients increase the risk of MDR-TB by 22.183 times [OR= 22.183,95% CI,1.829-268.12]. several studies documented close contact with DR-TB as the major risk factors of MDR-TB(37,41,42), which is contrast to the study conducted in Serbia (43).Our study showed that experience of hospital stays as a risk factor of MDR-TB [p = 0.001] ie, patient who had hospitalization history were more likely to develop MDR-TB than those who do not have previous history of hospital stay [p < 0.001, OR= 19.047, 95% CI, 7.92-45.76], which is also documented in other studies(25,44). The study conducted by v. Crudu et al documented that,in 75% of cases, the MDR-TB strain was genetically distinct from the non-MDR-TB strain at baseline, suggesting a high rate of nosocomial transmission of MDR-TB(40). Proper infection control measures in health facilities are crucial to protect other patients and healthcare workers, whereas preventive measures to reduce transmission within community is required.

This study showed that not attending TB counselling sessions [p < 0.001, OR = 5, 95%CI 2.92-10.90] is the risk factor for MDR-TB. One of the studies concluded that the Provision of counselling and financial support may not only reduce their vulnerability but also increase cure rates of MDR-TB(45). This study showed that smokers were more likely to develop MDR-TB than non-smokers [p<0.001, OR=4.8, 95%CI 2.34-10.11], as confirmed by other studies conducted in a different setting, china (46,47), Ethiopia(48), Lima Peru(49), Sudan(50), which is a contrast to the study conducted in Addis Ababa (36), Nepal(51). Smoking had also affected the global TB treatment success rate as shown in the study[65]. This study showed illicit drug use as the risk factor of MDR-TB [P = 0.002, OR = 8.33, 95%CI 1.82-38.06], which is in contrast to the study conducted in Canada(52) and Ethiopia(40). Engaging in illicit drug use and smoking can disrupt the balance of the immune system, potentially heightening susceptibility to infection.

This study showed that, stigma associated with TB as a risk factors of MDR-TB [p <0.001, OR=0.72, 95%CI 0.38-1.37], which is similar to the study conducted in Serbia(43), china(47), Nepal(45). It’s imperative to safeguard and defend the rights of all individuals with tuberculosis, as well as those most vulnerable to contracting the disease, Implementing policies and practices aimed at shielding them from stigmatization and discrimination is essential(4). There was significant difference in mental health of the respondents, this study showed Significant difference between the both groups in persistence of subjective feeling of nervousness [P < 0.001, OR=3.77, 95%CI 1.86-7.45], persistence of subjective feeling of sadness [p < 0.001, OR= 42.33, CI 1.36-1315.4] and experience of difficult situations in last 12 months [p < 0.001, OR=5.26 [2.416-11.488], which is similar to the study conducted in Serbia (43). The study conducted in china also showed experience more life pressure/stress [AOR, 10.8; 95% CI, 2.8–41.5] as the risk factors of MDR-TB(23)A life rejuvenation program tailored for tuberculosis patients in high-risk areas is imperative. As the study covers a large number of DR-TB cases in Nepal during the study period, the findings can be generalized.

## Strengths and Limitation

Strengths of the study include collecting information on recent episodes of previous TB treatment aimed at minimising recall bias. However, a notable limitation is the absence of data on additional treatment episodes, particularly concerning patients with multidrug-resistant tuberculosis [MDR-TB] who often undergo multiple treatments. Additionally, while the study successfully achieved its objectives, it did not employ multiple data-gathering methods to explore sensitive topics such as stigma and mental health issues. Notably, this study represents the first attempt to investigate the risk factors of multidrug-resistant tuberculosis in three districts of province number 3, which harbours the highest burden of DR-TB cases in Nepal, favouring generalisation of findings.

## CONCLUSION

This case-control study delved into factors associated with MDR-TB occurrence, identifying Pulmonary Tuberculosis, prior TB treatment history, close contact with DR-TB patients, and persistent subjective feelings of sadness as strong risk factors. The findings underscore the necessity for quality treatment coupled with attentive monitoring to curb drug resistance evolution among previously treated patients. Preventive measures should emphasize breaking transmission chains within communities, bolstered by robust infection control measures in healthcare facilities to minimize acquired MDR-TB cases among visitors, patients, and health workers. Additionally, the study underscores the pressing need for life rejuvenation programs targeting tuberculosis patients to safeguard their mental well-being.

## Data Availability

All relevant data are within the manuscripts table.

## List of abbreviations

AMR: Anti-Microbial Resistance
AOR: Adjusted Odds Ratio
BPKIHS: B.P. Koirala Institute of Health Sciences
CAT-I: Category I
CI: Confidence Interval
COPD: Chronic Obstructive Pulmonary Diseases
DOTS: Directly Observed Treatment Short Course
DR-TB: Drug-Resistant Tuberculosis
DST: Drug Susceptibility Test
DS-TB: Drug-Susceptible Tuberculosis
EPTB: Extra Pulmonary Tuberculosis
HF: Health Facility
HIV/AIDS: Human Immunodeficiency Virus/Acquired Immunodeficiency Syndrome
HR: Isoniazid, Rifampicin
HRZE: Isoniazid, Rifampicin, Pyrazinamide, Ethambutol
HRZELfx: Isoniazid, Rifampicin, Pyrazinamide, Ethambutol, Levofloxacin
JANTRA: Japan Nepal Health and Tuberculosis Research Association
LPA: Line Probe Assay
LR-1: Longer Regiment I
LR-2: Longer Regiment II
LR-3: Longer Regiment III
LTBI: Latent Tuberculosis Infection
M. Tuberculosis: Mycobacterium Tuberculosis
MDR-TB: Multi-Drug-Resistant Tuberculosis
MDR/RR-TB: Multi-Drug-Resistant Rifampicin-Resistant Tuberculosis
NATA: Nepal Anti Tuberculosis Association
NTC: National Tuberculosis Center
OR: Odds Ratio
PLS-PM: Partial Least Square Path Modelling
PTB: Pulmonary Tuberculosis
RR/MDR: Rifampicin-Resistant/Multi-Drug-Resistant
SDG: Sustainable Development Goals
SEAR: South East Asia Region
SPSS: Statistical Package for the Social Sciences
SSTR: Shorter Standardized Treatment Regimen
TB: Tuberculosis
TPT: Tuberculosis Preventive Therapy
TSR: Treatment Success Rate
WHO: World Health Organization
XDRTB: Extremely Drug-Resistant Tuberculosis

## Acknowledgements

The authors would like to acknowledge Dr Suvesh Kumar Shrestha for his valuable insights and sincerely thank all the participants.

## Author’s contribution

**Conceptualisation**: Puspa Acharya, Niraj Bhattarai, Vijay Kumar Khanal

**Data curation:** Puspa Acharya, Niraj Bhattarai, Dr Bhuban Raj Kunwar,Birendra Kuma Yadav,

**Formal analysis:** Puspa Acharya, Niraj Bhattarai, Khem Raj Sharma Methodology: Puspa Acharya, Niraj Bhattarai, Dr Bhuban Raj Kunwar

**Supervision:** Vijay Kumar Khanal, Birendra Kuma Yadav, Khem Raj Sharma, Dr Bhuban Raj Kunwar

**Validation:** Vijay Kumar Khanal, Birendra Kuma Yadav, Khem Raj Sharma, Dr Bhuban Raj Kunwar

**Writing original draft**: Puspa Acharya, Niraj Bhattarai

**Writing-review and editing:** Puspa Acharya, Niraj Bhattarai, Dr Bhuban Raj Kunwar

## Ethics

The study was conducted from March 2021 to August 2021. Ethical approval was obtained from the Institutional Review Committee [IRC] of BP Koirala Institute of Health Sciences [BPKIHS], Dharan, dated 18 Feb 2021, with IRC No. Acd/597. An original copy of the ethical approval is submitted as supplementary files. Verbal consent was obtained from the participants, and the study maintained their anonymity.

## Supporting Information

S 1: Original copy of ethical approval from the Institutional Review Committee.

